# COVID-19 In Shang Hai: It is Worth Learning from the Successful Experience in Preventing and Controlling the Overseas Epidemic Situation

**DOI:** 10.1101/2020.05.13.20100164

**Authors:** Qi Dang, Rui Miao, Yong Liang

**Affiliations:** Faculty of Information Technology, Macau University of Science and Technology, Avenida Wai Long, Taipa, Macau, China; State Key Laboratory of Quality Research in Chinese Medicines, Macau University of Science and Technology, Avenida Wai Long, Taipa, Macau, China; Guangdong-HongKong-Macao Joint Laboratory for Smart Discrete Manufacturing, Guangzhou 510006, China

**Keywords:** COVID-19, overseas imports, Shang Hai, Gaussian sparse network model, regression mode

## Abstract

COVID-19 first appeared in Wuhan, Hubei Province, China in late December 2019 and spread rapidly in China. Currently, the spread of local epidemics has been basically blocked. The import of overseas epidemics has become the main form of growth in China’s new epidemic. As an important international transportation hub in China, Shanghai is one of the regions with the highest risk of imported cases abroad. Due to imported of overseas cases are affected by the international epidemic trend. The traditional infectious disease model is difficult to accurately predict the cumulative trend of cumulative cases in the Shanghai areas. It is also difficult to accurately evaluate the effectiveness of the international traffic blockade. In this situation, this study takes Shanghai as an example to propose a new type of infectious disease prediction model. The model first uses the sparse graph model to analyze the international epidemic spread network to find countries and regions related to Shanghai. Next, multiple regression models were used to fit the existing COV-19 growth data in Shanghai. Finally, the model can predict the growth curve of Shanghai’s epidemic without blocking traffic. The results show that the control measures taken by Shanghai are very effective. At present, more and more countries and regions will face the current situation in Shanghai. We recommend that other countries and regions learn from Shanghai’s successful experience in preventing overseas imports in order to fully prepare for epidemic prevention and control.

## I. Introduction

SINCE December 2019, COVID-19 broke out in Wuhan, Hubei Province, China. Coronavirus disease (COVID-19) caused by the severe acute respiratory syndrome coronavirus 2 (SARS-CoV-2). this virus quickly spread throughout China. In the early stages of the COVID-19 epidemic, the number of infected cases increased exponentially[1]. The Chinese government has taken various preventive and control measures to intervene, including mandatory blockades, staggered working hours, etc.[2–4]. These measures effectively prevent the virus from spreading from person to person, and reduce the morbidity and mortality[5]. After more than two months, COVID-19 was gradually control in China and entered a stable state.

In view of the fact that the COVID-19 epidemic in China has been basically contained, other international regions of the world are still experiencing outbreaks. Unless substantive public health interventions are implemented, overseas countries and regions that have close communications with China may cause the epidemic in China to escalate again. Among them, Shanghai, as one of China’s largest international airports, is the region with the highest risk of overseas imports. Overseas case importation poses a huge threat to this city with a population of 24 million. As of 24:00 on April 2, 2020, there were 1562 confirmed cases in China, including 698 confirmed cases imported from abroad. The number of confirmed cases in Shanghai is 177, including 171 imported cases abroad. Therefore, in order to solve this problem. The Shanghai government has taken a lot of measures, including isolation observation, traffic control, and nucleic acid detection, etc. It is worth noting that from March 28th, Shanghai suspended the entry permit for non-Chinese nationals.

We also note that apart from Shanghai, many other regions in the world also face the same problem as Shanghai, that is, what measures should use to control the import of overseas cases? Since the outbreak of Wuhan, several modeling research teams around the world have used an infection model based on the SIR or SEIR framework to estimate and predict COVID-19[6–8]. However, the existing models are difficult to be directly applied to the prediction of overseas case trends. In this article, we propose a sparse relationship graph model for the establishment of an international epidemic spread network. After combining the regression model, we can predict the growth trend of cases in the target area. Because Shanghai, China is one of the earliest areas to take measures to block overseas spread. Therefore, this article takes Shanghai as an example, on the one hand, we want to verify the model proposed in this paper. On the other hand, we want to analyze whether the epidemic prevention and control measures taken by Shanghai are effective and whether Shanghai’s measures can provide an effective reference for similar regions.

We used 77-day real diagnosis data of 188 countries and regions around the world, and established an international network related to Shanghai. we discover that the epidemic trend in Shanghai is highly correlated with the epidemic situation in 30 countries and regions and most of these countries and regions are the center of the outbreak of the international epidemic. We established a variety of regression models to fit the existing growth curve, and predicted the growth of Shanghai cases from April 4th to 19th without blocking international traffic. This study believes that Shanghai’s blockade measures are necessary and effective, which avoids greater losses and effectively reduces the risk of secondary epidemics for China. We suggest that other countries learn from Shanghai’s successful experience and prepare well in advance when the next battle arrives.

## II. Materials and Method

### A. data source

The data in this article comes from the Johns Hopkins University Repository, mainly from January 19 to April 19, 2020, the number of daily diagnoses in various countries and regions around the world.

### B. Model

#### 1) Model of international relations network based on ***L_1_***

In order to discover the international correlation network of COVID-19, we build a sparse network model based on regularization, As the name implies, the network obtained by this model is a directionless network. The typical characteristic of a directionless network is that the degree between most nodes in the network is very small, the few nodes with large existence are called central points. For this study, our main focus is on nodes related to Shanghai, In the relationship network, if the model thinks that a country or region is related to Shanghai, it means that the epidemic situation in that country can affect the growth of cases in Shanghai. The mathematical framework of the sparse network model based on regularization in this paper is as follows[9]:

Consider the n-dimensional multivariate normal distributed random variable Equation (1):

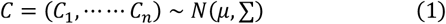

This includes Gaussian linear models. For example, *C*_1_ is the response variable,{*C_k_*; 2 ≦ *k* ≦ *n*} is a predictor, Assuming that the covariance matrix is nonsingular, you can use the graphical model *G* = (Γ, E) conveniently express the conditional independence structure of the distribution, In Γ = {1, ⋯ ⋯ *n*} is the set of nodes, and E is the set N of edges in C. Given all remaining variables *CΓ*\{a, b} = {*C_k_; k* ∈ *Γ*/{*a, b*}}, A pair of (*a, b*) is included in the edge set E if and only if C_B_ depends conditionally on C_C_. Given all remaining variables, each pair of variables not included in the edge set is conditionally independent and corresponds to the zero term in the inverse covariance matrix[10].

Neighborhood selection is a sub-problem of covariance selection. The minimum subset of the neighborhood Γ\{*a*} of the node *a∊*Γ, therefore, considering all the variables *C_a_* in the neighborhood, *C_a_* is conditionally independent of all remaining variables. The neighborhood of node*a∊*Γ consists of all nodes *a∊Γ*\{*a*;}, so ((*a*;, *b*)*∈*E. For the observation of C, neighborhood selection aims to estimate (individually) the neighborhood of any given variable (or node). Neighborhood selection can be used as a standard regression problem, it can be effectively solved with *L*_1_ [11], As will be shown in this paper.

For sparse high-dimensional graphs, the consistency of the proposed neighborhood selection will be shown, where the number of variables may grow with any power of the number of observations (high-dimensional), whereas the number of neighbors of any variable is growing at most slightly slower than the number of observations (sparsity).

Neighborhood selection with the *L*_1_. It is well known that the Lasso, introduced by Tibshirani [8], and known as Basis Pursuit in the context of wavelet regression[12], With simplicity [10]. When the forecast has all remaining variables *C_a_*{*C_k_; k∊*Γ\(*n*)\{*a*}}, The estimated value of the disappeared lasso coefficient asymptotically identifies the neighborhood of node *a* in the graph, as shown below. Let *n* × *c*_(*n*)_ – dimensional matrix *C* contain *c* independent *n* observations, so for all *a∊*Γ(*n*), column *C_a_* corresponds to a vector of *n* independent observations. Let〈·, ·〉 on ℝ_*n*_ be the usual inner product, and ‖·‖_2_the corresponding norm.

The *L*_1_ estimate *θ^a,λ^* of *θ^a^* is given by Equation (2):

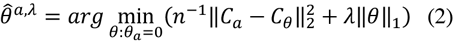

‖θ‖_1_ = ∑ *b∊*Γ_(*n*)_ |θ_*b*_| is the ***L*_1_**-norm of the coefficient vector. It is recommended to normalize all variables to a common empirical variance in the above formula. The solution of the above formula is not necessarily unique. However, if the uniqueness fails, the solution set is still convex, and all of our results on the neighborhood apply to any solution of the above formula.

Other regression estimates based on the *lp* norm have been proposed, where p is usually in the range [0, 2] (see[13]). A value of *p* = 2 will result in ridge estimation, while *p* = 0 corresponds to traditional model selection. As we all know, only when *p* ≤ 1, the estimated value has a parsimony property (some components happen to be zero), For *p* ≥ 1, the optimization problem in the above formula is only convex. Therefore, the minimization of empirical risk constrained by ***L*_1_** occupies *a* unique position, because *p* = 1 is the only value of *p*, variable selection is made at this value, and the optimization problem is still convex, so it is feasible for high-dimensional problems.

The neighborhood estimate (parameterized by *λ*) is defined by the non-zero coefficient estimate of *L*_1_ penalty regression Equation (3):

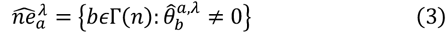

Therefore, each choice of penalty parameter *λ* specifies an estimate of the neighborhood of node *a∊*Γ(*n*), and the rest is to choose the appropriate penalty parameter. A larger penalty value tends to reduce the size of the estimated set, and if the value of *λ* decreases, usually more variables are included in the estimated value.

The prediction-oracle solution. A seemingly useful choice of penalty parameters is (unavailable) to predict the oracle value Equation (4):

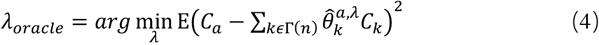

Expectation is understood to be abo has nothing to do with samples that estimate *θ^a,λ^*. The prediction penalty minimizes the prediction risk in all L_1_ regular sub-estimates. The *λoracle* estimate is obtained by selecting *λcv* for cross-validation.

Shao[14] showed that for *L*_0_ penalty regression, the cross-validation choice of penalty parameters is consistent with the model choice of the verification set size under certain conditions. Predicting that the Oracle solution will not lead to consistent model selection for *L*_1_, as shown in the simple example below.

Proposition. Let the number of variables grow to infinity, for *n* → ∞, *p*(*n*) → ∞, for some γ > 0 with *p*(*n*) = *o*(*n^γ^*), Assume that the covariance matrices ∑(*n*) except for some pair (*a, b*)*∊*Γ(*n*) × Γ(*n*), for which ∑_*ab*_(*n*) = ∑_*ba*_(*n*) = *s*, for some 0 < *s* < 1 and all *n∊N*_o_ under the prediction-oracle penalty, The probability of choosing the wrong neighborhood for node *a* converges to 1, as shown in Equation(5):

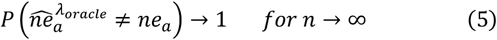

From the proof of the above proposition, it can be concluded that many noise variables are included in the prediction of the neighborhood of the Oracle solution. In fact, for a fixed number of variables, the possibility of including noise variables in prediction predictions will not even disappear. If the selected penalty is greater than the predicted optimal value, then the *L*_1_ regularizer can be used for consistent neighborhood selection.

#### 2) Regression model

After establishing a global epidemic spread relationship network and identifying relevant countries and regions in Shanghai. In this paper, three regression models are established for experiments including: nuclear ridge regression model, elastic network model and Bayesian regression model.

##### a) Kernel Ridge Regression

For the parameter matrix, in order to solve the risk of overfitting, we sequentially add “kernel” and “ridge” to the linear regression model to transform the model[15].

The first step is to add “kernel”. For a given test sample, each training sample 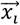 contains a coefficient *α_j_*, so the loss function can be expanded as shown in Equation (6).

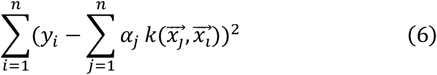

This is a quadratic loss function. We choose the appropriate *α_j_* to maximize it.

The second step, adding “ridge”, is actually adding regularization terms, adding an L_N_ regularization on the basis of linear regression. Then the loss function becomes shown in Equation (7).

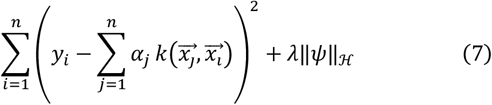

The second summation part of the above formula will penalize large coefficients, which can prevent the model from giving a large weight to a single training example and overemphasizing the role of a single training example, the parameter λ controls the trade-off between the degree of fitting and the complexity of the model.ℋis Hilbert space, ‖∙‖_ℋ_ is a 2-paradigm under Hilbert space. Expanding the regularization term, the loss function can be changed to the form shown in Equation (8).

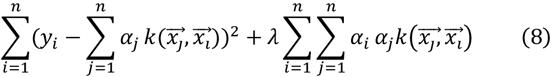

Among them, K is a matrix of 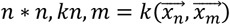. 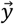 is a vector of n labels.

##### b) Elastic network

Elastic network is a linear regression model trained using ***L*_1_** and ***L*_2_** norms as a priori regular terms[16]. This model can fit non-zero sparse matrices with a small number of parameters. Elastic networks are very useful when many features are interrelated. Elastic networks are more inclined to consider two of these characteristics randomly, and inherit the stability of Ridge during the allowed cycle[17].

Here, the objective function of minimization is shown in Equation (9).

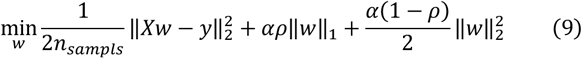

##### c) Bayesian Ridge Regression

Bayesian Ridge Regression is to use the probability model to estimate the regression model, mainly to solve the complexity of the model in the process of maximum likelihood estimation[18]. The process of Bayesian Ridge regression is a process in which sample points are gradually added to the learner. The posterior of the previous sample point will be used as a priori by the next estimation. In other words, Bayesian learning is gradually updating the prior. The prior update is actually iterated by updating the maximum likelihood estimation parameter and sample points, the prior parameter ω is obtained by the Gaussian mode Equation (10).

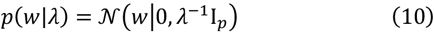

The prior parameters α and λ generally follow the gamma distribution, and this distribution has a conjugate prior relationship with Gaussian [19]. The parameters ω,;α and λ are estimated together when the model is fitted, where the parameters α and λ are obtained by maximum likelihood estimation.

The difference between Bayesian Ridge Regression and ordinary Ridge Regression is that it adopts Bayesian strategy of updating priors’ step by step. Ordinary Ridge Regression allows the parameter to be zero, but Bayesian Estimation cannot do so because the standard deviation of Gaussian Can be infinite. At the same time, Bayesian regression will give the confidence interval of the parameter, which is an optional range of the parameter, which is essentially a covariance matrix.

## III. Experiment

This chapter first uses the case data of 188 countries and regions in the world from January 19 to April 4 to accumulate a total of 77 days to establish a global epidemic transmission relationship network. Next, the subnet data of countries related to Shanghai are extracted. Then, three regression models were used to establish regression models to fit the cumulative case growth data from January 19 to April 4 in Shanghai. This article mainly uses the 5-fold cross-validation, explained_variance, mean_absolute_error, mean_squared_error and r2 indicators to verify the model results. Finally, through the regression model after training, we predicted the cumulative diagnosed cases in Shanghai from April 4 to April 19(without blocking traffic).

### A. Global network analysis

According to the latest China national epidemic prevention and control instructions “anti-import, non-proliferation”, the Shanghai government responded to the Chinese government’s call. On March 28, 2020, the entry application for non-Chinese nationals was stopped. The infection period of COVID-19 seems to be very long, which may last 10 days or longer after the incubation period [20]. Considering the special situation, we chose a latency of 7 days. Therefore, through the sparse network model based on L_1_, we constructed the data matrix of the actual case statistics of 188 countries and regions in the world from January 19th to April 4th, and plotted the COVID-19 international network. As shown in Figure 1, by analyzing the international relations network, we have counted the central nodes of the international epidemic transmission network, As shown in Table 1, we found that the United States, Italy, Iran, and Hubei, China are the main Hub nodes, that is, the main international epidemic spread countries. This result is consistent with the existing real situation; Therefore, it can be concluded that the international network relations based on ***L_1_*** in this study have certain reliability.

**Figure. 1.**
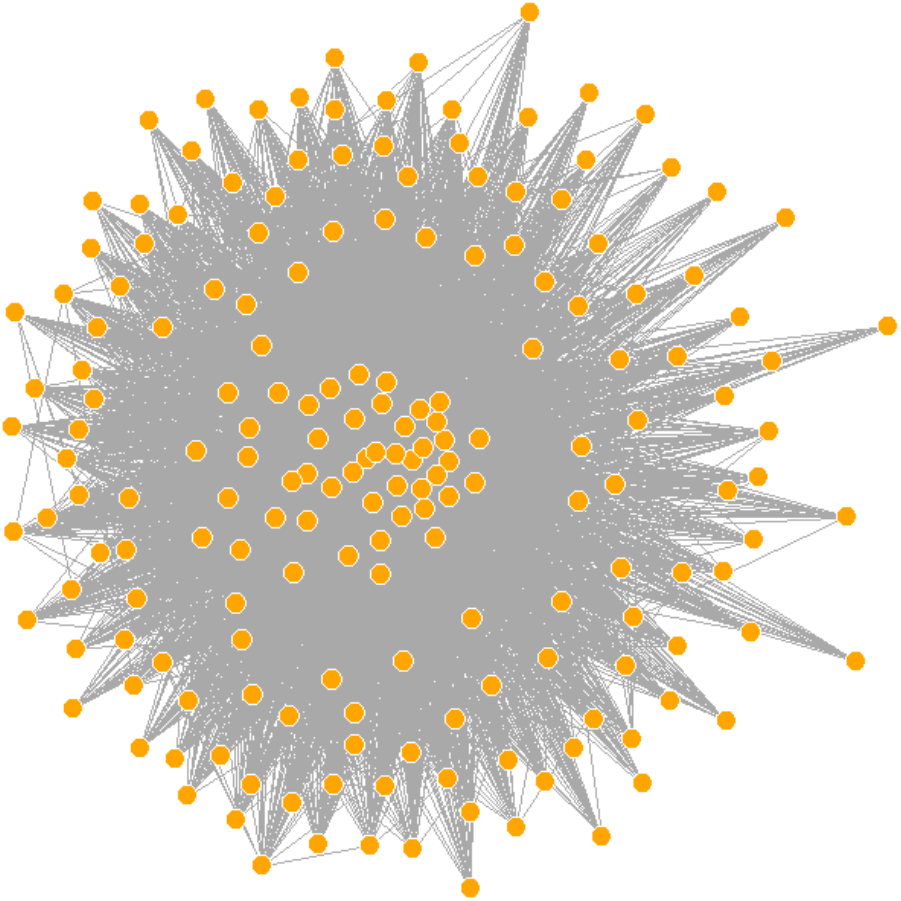
COVID-19 International Relations Network in 188 countries and regions

**Table I.** Hub nodes in the international epidemic relationship network

| Country | United States | France | Germany | Italy | Iran | Hubei, China |
| --- | --- | --- | --- | --- | --- | --- |
| Number of nodes | 183 | 183 | 183 | 181 | 180 | 180 |

### B. Shanghai Subnet Analysis

Next, we extracted the relationship subnets of cities and regions related to Shanghai. as shown in Figure 2. We can see that Shanghai is closely related to many areas of China, such as Hubei, Guangdong, Beijing and Heilongjiang. This is because in the early days of the spread of the Shanghai epidemic, these areas were the main import areas to Shanghai. At present, the epidemic situation in these areas has been stable and basically contained. As shown in Figure 2, we can also find many international communication channels, such as: The United States, European countries, Iran, the Philippines and other regions, this is caused by COVID-19 broke out in the international scope, brought by passengers entering international flights Input abroad. After identifying 30 countries and regions related to Shanghai, we can further establish a regression model to fit the epidemic growth data of Shanghai from January 19 to April 4.

**Figure. 2.**
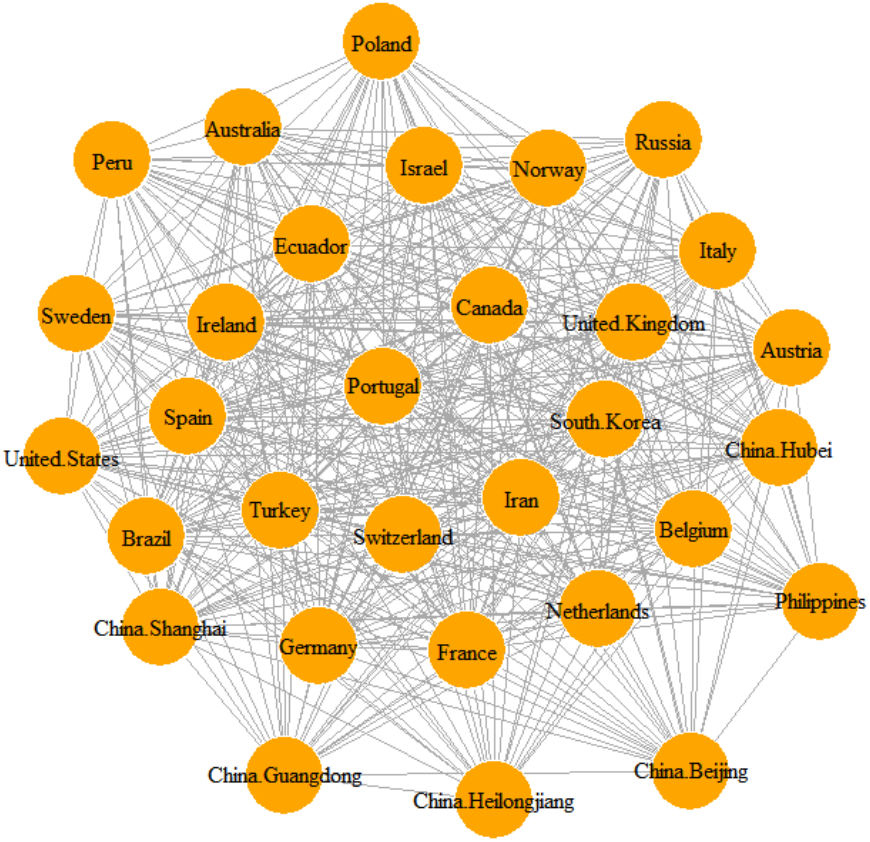
COVID-19 International Relations Network in 30 countries and regions related to Shang Hai

### C. Building a regression model

This article uses three regression algorithms to build the model, which are: KernelRidge, BayesianRidge and ElasticNet model. As shown in Figure 3 and Table 2–Table 3, the three methods can fit the existing growth data in Shanghai well. Among them, the ElasticNet model performs best, explained_variance, mean_absolute_error, mean_squared_error and r2 values Only 0.997, 3.19, 29.68 and 0.997, respectively, the average loss value of the 5-fold cross-validation is only –6.624. It can also be seen from the fitting curve that this method shows a good fitting effect. Experiments have proved that without blocking international traffic in Shanghai, The real data of the countries and regions related to Shanghai in the above international relations networkCan effectively predict the growth of cases in Shanghai.

**Figure. 3.**
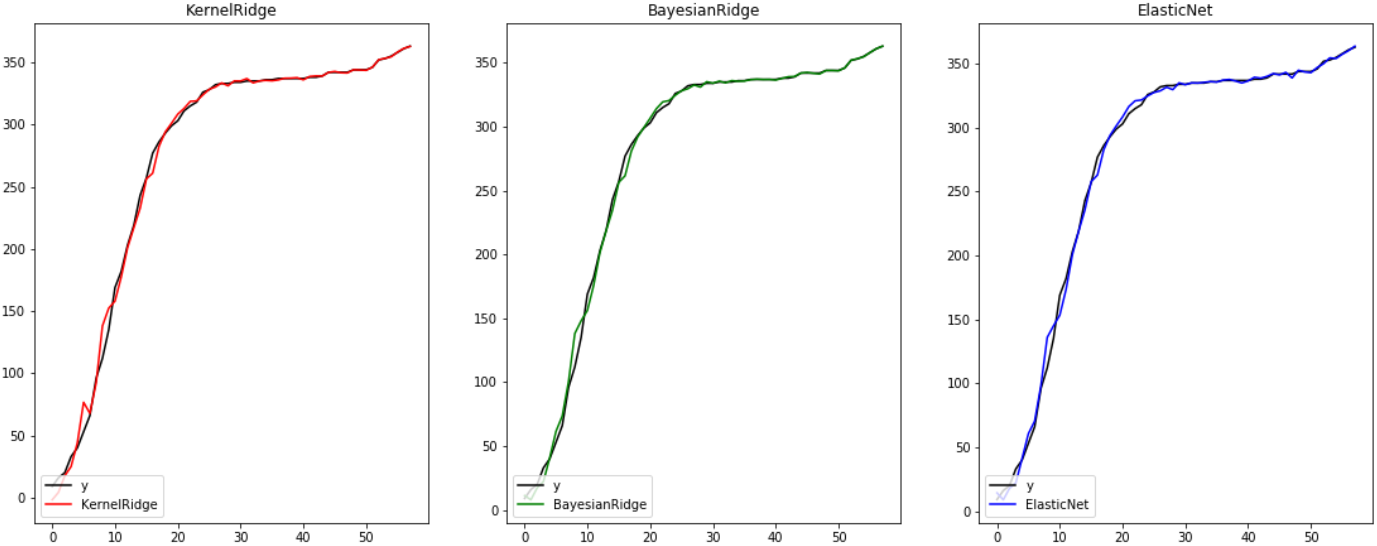
Comparison of cumulative confirmed cases and actual values predicted by the three regression methods

**Table II.** The results of three regression methods based on 5-fold verification

|  | 0 | 1 | 3 | 4 | 5 |
| --- | --- | --- | --- | --- | --- |
| KernelRidge | -7.8 | 0.62 | -11.51 | -12.12 | -18.64 |
| BayesianRidge | -12.14 | -2.6 | -21.18 | -7.6 | 24.04 |
| ElasticNet | -10.68 | 0.92 | -5.41 | -0.17 | -22.78 |

**Table III.** Performance of different indicators of the three regression models

|  | ev | mae | mse | r2 |
| --- | --- | --- | --- | --- |
| KernelRidge | 0.996 | 3.39 | 43.12 | 0.996 |
| BayesianRidge | 0.997 | 2.82 | 31.10 | 0.997 |
| ElasticNet | 0.997 | 3.19 | 29.68 | 0.997 |

### D. Short-term epidemic growth curve (unblocked international traffic)

After establishing a regression model, we use data from 30 countries and regions related to Shanghai from April 4 to April 19 for 15 days to predict Shanghai’s epidemic growth curve without the international traffic blockade, and compare it with real data. The experimental results are shown in Figure 4 and Table 4. On the premise that international traffic is not blocked, all three regression models have concluded that the cumulative number of confirmed cases will far exceed the existing real data. On April 19th, the cumulative case prediction results of the three models were between 1154–1386 people. In fact, the cumulative confirmed cases on that day were 526, and the predicted data was slightly greater than twice the true confirmed cases. The result of this comparison shows that Shanghai’s adoption of the international traffic blockade has a significant effect and can largely prevent the import of overseas epidemic situations. It also means that Shanghai’s traffic control is effective and necessary.

**Figure 4.**
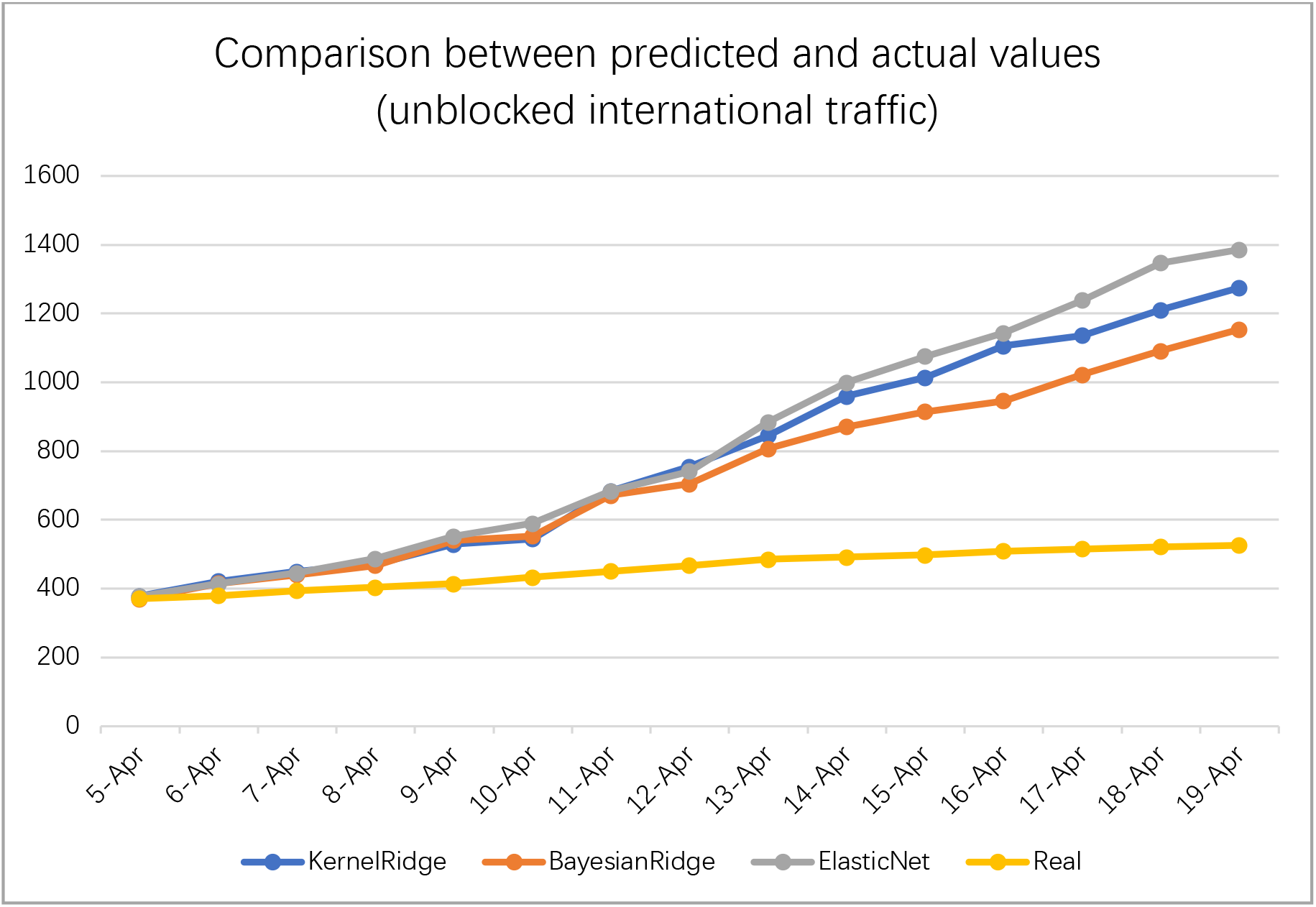
Comparison between predicted and actual values (unblocked international traffic)

**Table IV.** Comparison between predicted and actual values (unblocked international traffic)

|  | KernelRidge | BayesianRidge | ElasticNet | Real |
| --- | --- | --- | --- | --- |
| April 5 | 379 | 370 | 378 | 371 |
| April 6 | 422 | 415 | 414 | 380 |
| April 7 | 450 | 440 | 446 | 394 |
| April 8 | 471 | 468 | 487 | 404 |
| April 9 | 529 | 541 | 552 | 414 |
| April 10 | 545 | 553 | 590 | 433 |
| April 11 | 684 | 671 | 684 | 451 |
| April 12 | 755 | 705 | 741 | 468 |
| April 13 | 846 | 807 | 884 | 485 |
| April 14 | 960 | 871 | 999 | 492 |
| April 15 | 1014 | 915 | 1076 | 498 |
| April 16 | 1106 | 946 | 1144 | 509 |
| April 17 | 1136 | 1022 | 1239 | 516 |
| April 18 | 1211 | 1091 | 1347 | 522 |
| April 19 | 1275 | 1154 | 1386 | 526 |

## IV. Discussion

COVID-19 quickly spread from a city to the entire country in just 30 days. This alarming rate of expansion and the number of cases growing shocked the public health service system of the entire country. The Chinese government showed to the world its epidemic response capacity in the first time. These measures have made a significant effect in responding to COVID-19. China’s domestic epidemic situation has been basically blocked. At present, COVID-19 has broken out in many countries outside the country. Therefore, at this stage, the main way of confirmed cases in China is passengers entering by international transportation.

This article establishes a network of epidemic transmission relationships between Shanghai and different countries and regions around the world, and builds a regression model based on network information to fit Shanghai’s COVID-19 epidemic growth data. The results show that the regression model based on the relational network can better fit the existing cumulative number growth curve. Combined with the regression model, we can predict the future development trend of cumulative cases in Shanghai (unblocked international traffic). According to the prediction results, we learned that the Shanghai government adopted traffic blockade measures, which effectively prevented the epidemic from spreading further. At present, the imported epidemic situation outside Shanghai has been effectively controlled, but we still need to be vigilant to prevent the recurrence of the epidemic situation. More importantly, with the effective control of the local epidemic situation in various countries, Shanghai’s successful experience in dealing with the import of overseas epidemic situations has also played a demonstration role for other countries and regions.

In order to reduce the risk of imported COVID-19 cases, Shanghai has adopted a series of measures to prevent and control possible overseas imported cases, strengthen monitoring and rapid identification of possible cases. First, during the epidemic, all Chinese and foreign personnel who have lived in key countries or regions within 14 days before entering Shanghai will be quarantined for 14 days. Secondly, conduct temperature examination and medical observation on all entry personnel, and then transfer and isolate confirmed cases and suspected cases for rapid treatment. Carry out preventive measures for close contacts. Since March 28, international routes have been suspended, and foreign tourists are prohibited from entering Shanghai.

As a successful import case for overseas epidemic prevention, Shanghai has fully demonstrated how a region can prevent a global pandemic, and it has provided a good demonstration for other countries and regions. At the same time, it also showed the friendly style of the Chinese nation and treated all immigrants equally during the epidemic, making foreign friends feel warm during this difficult time.

## ACKNOWLEDGMENT

This work is supported by the Macau Science and Technology Development Funds Grands No.003/2016/AFJ and No.0055/2018/A2 from the Macau Special Administrative Region of the People’s Republic of China.

